# Bidirectional genetic and phenotypic links between smoking and striatal iron content involving dopaminergic and inflammatory pathways

**DOI:** 10.1101/2024.09.26.24314454

**Authors:** Olga Trofimova, Ilaria Iuliani, Sven Bergmann

## Abstract

Tobacco smoking is a major modifiable risk factor for cardiovascular and lung diseases. A better understanding of its neurobiological underpinnings will benefit the prevention of smoking-related illnesses and mortality. Recent neuroimaging studies have identified a correlation between smoking and iron concentration in the brain’s striatum, a subcortical region involved in habit formation and compulsive behaviour, and a central node of dopamine activity. Moreover, iron accumulation in the striatum is associated with lower cognitive performance in adults. Here, we investigated phenotypic and genetic correlations, and causal relationships between smoking initiation (ever smoked regularly) and susceptibility-weighted magnetic resonance imaging (MRI)-derived markers of iron content–T2* and quantitative susceptibility mapping (QSM)–in the bilateral putamen, caudate, and accumbens nuclei. We computed correlations between smoking and striatal iron in the UK Biobank, adjusting for a vast set of imaging and non-imaging confounders. Using genome-wide association studies (GWAS) summary statistics, we performed global genetic correlation, cross-GWAS coherence tests at the gene level, and causality analysis using Mendelian randomisation and PascalX. Smoking was positively correlated with iron content in the bilateral putamen, caudate, and in the left accumbens, with the strongest effect found when contrasting current and never smokers. Striatal iron had a positive association with pack-years and a negative relationship with years since stopping smoking, indicating a possible reversal of iron accumulation after smoking cessation. Genetic correlation paralleled phenotypic correlation. Cross-GWAS signal was coherent in genes involved in the dopaminergic and glutamatergic systems, and synaptic function. There was evidence of a causal relationship from smoking to striatal iron through genes involved in synaptogenesis and plasticity, and to a lesser extent, from striatal iron to smoking through inflammatory and immune system related genes. Moreover, the heterogeneity of genes with correlated and anti-correlated signals suggests that the neurobiological mechanisms linking iron to smoking behaviour are highly complex. Overall our results show an association between cigarette smoking and iron concentration in the striatum with complex multi-directional causal mechanisms involving synaptic transmission and inflammatory circuits.

## Introduction

Smoking is a known risk factor for many diseases, including lung cancer, cardiovascular disease, and respiratory disorders. The enormous healthcare costs associated with the adverse effects of tobacco smoking make it a major public health concern. While the effects of smoking on the lungs, cardiovascular system, and cancer risk have been extensively studied (1–3) its impact on the brain has received less attention.

Existing literature primarily focuses on white matter ageing (4–6) and grey matter volumetry (4,5,7–13). Other studies have examined the relationship between smoking, cerebrospinal fluid biomarkers, and cognitive impairment (14,15). A few studies have reported increased iron concentration in the striatum of smokers (16–18).

The striatum is central to motivation, reward, habit formation, addiction, and compulsive behaviour, with dopamine and glutamate being the predominant neurotransmitters involved (19). Iron accumulation in the striatum is associated with lower cognitive performance in older age and increased vulnerability to brain injury from stroke (20). Additionally, iron accumulation is a common feature of most neurodegenerative diseases (20–22). Processes such as ferroptosis, an iron-dependent form of cell death driven by lipid peroxidation (22,23), and iron-induced neuroinflammation are believed to play roles in Alzheimer’s disease and vascular dementia (24). More generally, iron is considered an indirect marker of oxidative stress (21,25,26).

Given the potential negative clinical consequences of brain iron overload, an important question arises: is there a causal relationship between smoking and iron accumulation in the brain? Existing Mendelian randomisation (MR) studies on alcohol and iron (27), smoking and white matter (6), or subcortical volumes and smoking and alcohol (10) tend to find that behaviour causes changes in brain structure rather than the other way around. Understanding the biological mechanisms at play could provide crucial insights for clinicians treating nicotine addiction.

In this study, we explored the relationship between smoking and striatal iron accumulation at the phenotypic and genetic levels, investigating both the potential causality and underlying biological mechanisms. Given the role of iron in dopamine synthesis and signalling (22,26,28,29) and the neuroimaging-based evidence for dopamine increase in the striatum following cigarette smoking (30), we hypothesised that dopamine-related genes would be jointly involved in smoking and striatal iron. We found a positive association between smoking and iron content in the dorsal striatum, with evidence suggesting that smoking may primarily drive iron accumulation rather than the reverse. Additionally, we identified complex, bidirectional genetic relationships involving dopamine-related genes, along with genes linked to synaptic function, GABAergic and glutamatergic transmission, inflammatory response, and the immune system.

## Materials and Methods

### Participants

We conducted our study on data from the UK Biobank (UKB), a large-scale biomedical database that comprises extensive health and genetic information from over 500k participants aged between 40 and 69 years at the time of recruitment (2006–2010) (31). We included participants with available brain imaging data and excluded those for whom smoking status was missing.

### Brain imaging

The UKB brain imaging sub-sample included approximately 40k participants aged from 44 to 85 years at the time of image acquisition (16). Our study made use of imaging-derived phenotypes (IDP) from susceptibility-weighted magnetic resonance imaging (swMRI) generated by an image-processing pipeline developed and run on behalf of UKB (32). More specifically, we examined median T2* and median quantitative susceptibility mapping (QSM) in bilateral putamen, caudate, and accumbens nuclei, corresponding to UKB data-fields (DF) 24469-24472, 24479, 24480, 25028-25031, 25038, and 25039. T2* and QSM are swMRI techniques used to assess tissue iron content, with T2* providing a measure of signal decay influenced by iron deposits and QSM offering a quantitative map of tissue magnetic susceptibility to precisely quantify iron levels. Higher iron content results in lower T2* and higher QSM values. Both techniques are also influenced in different ways by other tissue properties, such as myelin, calcium, and water, making them complementary (33). For example, while myelin and iron have the same effect on T2*, they have opposite effects on QSM (18).

### Smoking

Smoking data was self-reported at the imaging visit. Smoking status (DF 20116) was categorised as current, former, or never having smoked. Pack-years of smoking (DF 20161) was defined as the number of cigarettes smoked per day divided by twenty and multiplied by the number of years of smoking. The latter was derived by subtracting the age of starting smoking from either the current age or the age at which smoking was stopped. ‘Years since stopping smoking’ (YSSS) was calculated as the participant’s age (DF 21003) minus the age at which they stopped smoking (DF 2897).

### Covariates

General covariates included age (DF 21003), age^2^, sex (DF 31), age ✕ sex, age^2^ ✕ sex, Townsend deprivation index (DF 22189), household income before tax (DF 738), educational attainment (highest among qualifications reported in DF 6138), alcohol intake frequency (DF 1558), diastolic and systolic blood pressures (mean of the two measurements in DFs 4079 and 4080), and body mass index (BMI, DF 21001). Imaging covariates (Alfaro-Almagro 2021) included assessment centre (DF 54), date (DF 53) and date^2^, protocol phase (DF 25780), protocol change affecting swMRI (DF 24418), head size (DF 25000), scanner and table position (DFs 25756-25759), head motion (DFs 24419, 24441, 24447, 24453), variations in acquisition protocols (DFs 25921, 25922, 25923, 25924, 26500) and intensity scalings (DFs 25925-25930). All covariates were collected at the imaging visit except sex and Townsend deprivation index, which were collected at the first visit. We considered ‘Do not know’ and ‘Prefer not to answer’ as missing values. We imputed missing covariate values by replacing them with the group mean for continuous variables or mode for categorical ones.

### Phenotypic association between smoking and striatal iron

First, we conducted some data preprocessing steps. We log-transformed pack-years of smoking and Townsend deprivation index due to skewness in the data. We excluded outlier IDP values beyond five absolute median deviations from the group median. At that stage, we examined brain IDP data distributions, compared male and female means with a two-sample t-test with Cohen’s d for effect sizes, male and female variances with an F-test, and computed Pearson’s correlations between age and IDPs. We also plotted IDP values stratified by smoking status and sex for visualisation purposes but without statistical testing, since this was investigated later using linear regression models, as described in the next paragraph. Next, on the brain IDPs and smoking variables, we applied a de-confounding procedure that consisted of regressing out the above-listed imaging and non-imaging covariates, to use the residuals as our new variables. We then performed a rank-based inverse normal transformation and z-scoring on continuously distributed variables (i.e. de-confounded IDPs, pack-years, and YSSS).

Next, we performed ordinary least squares linear regression models to analyse the relationship between the 12 IDPs–T2* and QSM in left and right putamen, caudate, and accumbens–and six smoking variables–ever smoked (current and former smokers against never smokers), currently smoking (current against former and never), current vs never (excluding former smokers), current vs former (excluding never-smokers), former vs never (excluding current smokers), and pack-years. In all models, the IDP was the response variable, and smoking was the predictor.

We also investigated the interaction effects of smoking with sex and age. For former smokers, we ran linear regression models with YSSS, pack-years, and their interaction term as predictors. For all phenotypic association analyses, we applied false discovery rate (FDR) correction for multiple (6 smoking ✕ 12 brain phenotypes) testing (34). We set the alpha threshold at 0.05 on FDR-corrected p-values. As a post-hoc analysis of the left-right asymmetry found in the accumbens QSM associations with the six smoking variables, we compared the respective beta coefficients from the left and right hemispheres using a z-test.

### Genetic correlation and causal relationship

We performed the genetic analyses using publicly available genome-wide association study (GWAS) summary statistics computed for UKB brain IDPs from ∼33k unrelated individuals with recent UK ancestry (18,35,36). For smoking, we utilised summary statistics from the larger GWAS and Sequencing Consortium of Alcohol and Nicotine use (GSCAN) study, which included the UKB and 29 other cohorts of European-ancestry individuals, for a total of ∼500k participants, excluding 23andMe for which GWAS summary statistics publication is not allowed (37). We chose ‘smoking initiation’ as our variable of interest, which was the equivalent of ‘ever smoked’ from the phenotypic analysis part of our study.

We computed the global genetic correlation between each of the 12 brain IDPs and smoking initiation using linkage disequilibrium score regression (LDSR) (38) across approximately 1.2M single nucleotide polymorphisms (SNPs). As for the phenotypic analysis, we applied FDR correction (34) to the 12 obtained p-values. We then used PascalX cross-GWAS coherence tests (39,40) to compute the gene-wise correlation between smoking and the IDPs. PascalX assesses coherent effects across the SNPs associated with two traits within a 50kb gene window, accounting for linkage disequilibrium (LD). We first investigated five dopamine-related candidate genes expressed in the striatum and previously associated with smoking initiation (*DRD1*, *DRD2*, *DRD3*, *DRD4*, and *PPP1R1B*) (29,37,41–44), and then an exhaustive list of 18 344 protein-coding genes. The LD structure required by PascalX to compute cross-GWAS coherence scores was provided with the UK10K (hg19) reference panel (45). Gene annotations were obtained from Ensembl, GRCh37 version (Ensembl release 75) (46). For positive correlations, we tested QSM coherence and T2* anti-coherence with smoking, while for negative correlations, we tested QSM anti-coherence and T2* coherence. Given the brain GWAS sample size, we included in the analysis all SNPs with a minor allele frequency of at least 0.01, which ensured we got a minimum of ∼33 individuals with at least one minor allele.

We mapped SNPs to genes according to the GRCh37 genome assembly (47). SNP p-values were rank-normalised prior to the cross-GWAS coherence scoring, making the test more conservative but less prone to type-1 errors. We also corrected for sample overlap using the LDSR intercept since the brain GWAS was nested in the smoking GWAS in terms of participants (40). We corrected for multiple testing with the Bonferroni method, i.e. by setting the alpha threshold to 0.05 divided by the number of genes (18 344 for the exhaustive list of genes and 5 for the candidate genes). When two or more genes located on the same chromosome, arm, and position displayed significant results, we report them here as gene clusters. In such cases, it was not possible to know what gene(s) was (were) driving the signal, given that the gene window we used (50kb) could be too large for smaller genes.

We investigated potential causal relationships between smoking and striatal iron content with two distinct techniques – Mendelian randomisation (MR) and our recently proposed PascalX cross-GWAS coherence ratio test.

MR is a powerful method commonly used to infer causal relationships between an exposure and an outcome using genetic variants as instrumental variables (IVs) (48). Here we performed bidirectional two-sample MR analyses using the TwoSampleMR R package (49). We selected independent SNPs significantly associated with the exposure (p < 5×10^-8^) as genetic instruments, pruning SNPs with r^2^ > 0.001 to a lead SNP according to LD estimates from the UK10K reference panel (45,50). When fewer than five IVs were available, we used a less stringent p-value threshold (p < 10^-5^). Causal estimates were computed using the inverse-variance weighted (IVW) method, and we applied FDR for multiple testing correction (34,51).

The PascalX ratio test (39) also employs the GWAS summary statistics from two traits. Its test statistic is computed for each gene by summing over the products of the respective effect sizes from all SNPs within a 50kb window around the gene’s transcribed region. Importantly, this sum is then normalised by the sum of squared effects from the outcome trait. Similar to MR, this test statistic can only deviate substantially from zero if the exposure and outcome effects are correlated with each other (either positively or negatively). The normalisation also ensures that the effects of the exposure that contribute to this correlation have to be large (resembling the MR requirement that the instrument variables need to be strongly associated with the exposure), while this is not the case for the output. Significant genes can then inform us upon the biological mechanisms underlying causal links, but results must be interpreted carefully when confounding effects from other variables cannot be excluded (for more details on the ratio test, see (39)). The PascalX parameters were the same as for the cross-GWAS coherence test, including the Bonferroni correction for the number of tested genes.

## Results

### Participant characteristics and striatal iron distribution by sex, age, and smoking status

The phenotypic analysis included 41 844 UKB participants, of whom 22 156 (52.9%) were female. The average age was 64.2 years (±7.7). Smoking status was distributed as follows: 3.3% were current smokers, 33.9% were former smokers, and 62.8% had never smoked (further covariate descriptive statistics can be found in Supp. Table 1).

Prior to data de-confounding and normalisation, we investigated T2* and QSM data distributions as a function of sex, age, and smoking status. Females had generally lower iron than males, as reflected by lower QSM and higher T2* values (except in the right accumbens), although the effect sizes were small (|Cohen’s d| ∈ [0.04, 0.24], see Supp. Fig. 1). The variance of IDPs did not differ between males and females (F values in Supp. Fig. 1). Iron concentration was higher in older participants in the putamen and caudate (|r| ∈ [0.11, 0.33]) but did not differ much in the accumbens (|r| ∈ [0.02, 0.08], Supp. Fig. 2). In both males and females, iron was generally higher in former smokers than in never smokers, and in current smokers than in former smokers (Supp. Fig. 3), although significance was not tested at that stage, given that the correlation between smoking and the IDPs was thoroughly investigated after variable de-confounding.

### Phenotypic association between smoking and striatal iron

Smoking was consistently associated with higher iron content in the bilateral putamen and caudate, as indicated by positive correlations with QSM and negative correlations with T2* (Fig. 1a and Supp. Table 2). The strongest effects (|β| ∈ [0.23, 0.4]) were observed when contrasting current against never smokers. In the accumbens, the association with smoking was only present when using QSM as a marker of iron and when contrasting current smokers against other groups, but not former against never smokers nor when measuring smoking in pack-years. Interestingly, the association signals in the accumbens were stronger in the left than in the right hemisphere, although the asymmetry was only significant for ‘ever smoked’ (z = 1.74, p = 0.04; see Supp. Table 3). Sex and age interactions with smoking variables were not significant, indicating that the strength of association between smoking and striatal iron IDPs was similar across ages and sexes (Supp. Fig. 4 and Supp. Table 2). Among former smokers, YSSS, pack-years, and their interaction were significant in the putamen and caudate but not in the accumbens (Fig. 1b and Supp. Table 4). This interaction effect is shown in Supplementary Figure 4, where the QSM intercept was higher in individuals with higher pack-years, the slope associated with YSSS was generally negative, and steeper for higher pack-years values. In other words, the more years passed since smoking cessation, the lower the dorsal striatal iron levels, and the smaller the difference in iron levels between heavy and light smokers. The same was observed with opposite signs in T2*.

**Figure 1.**
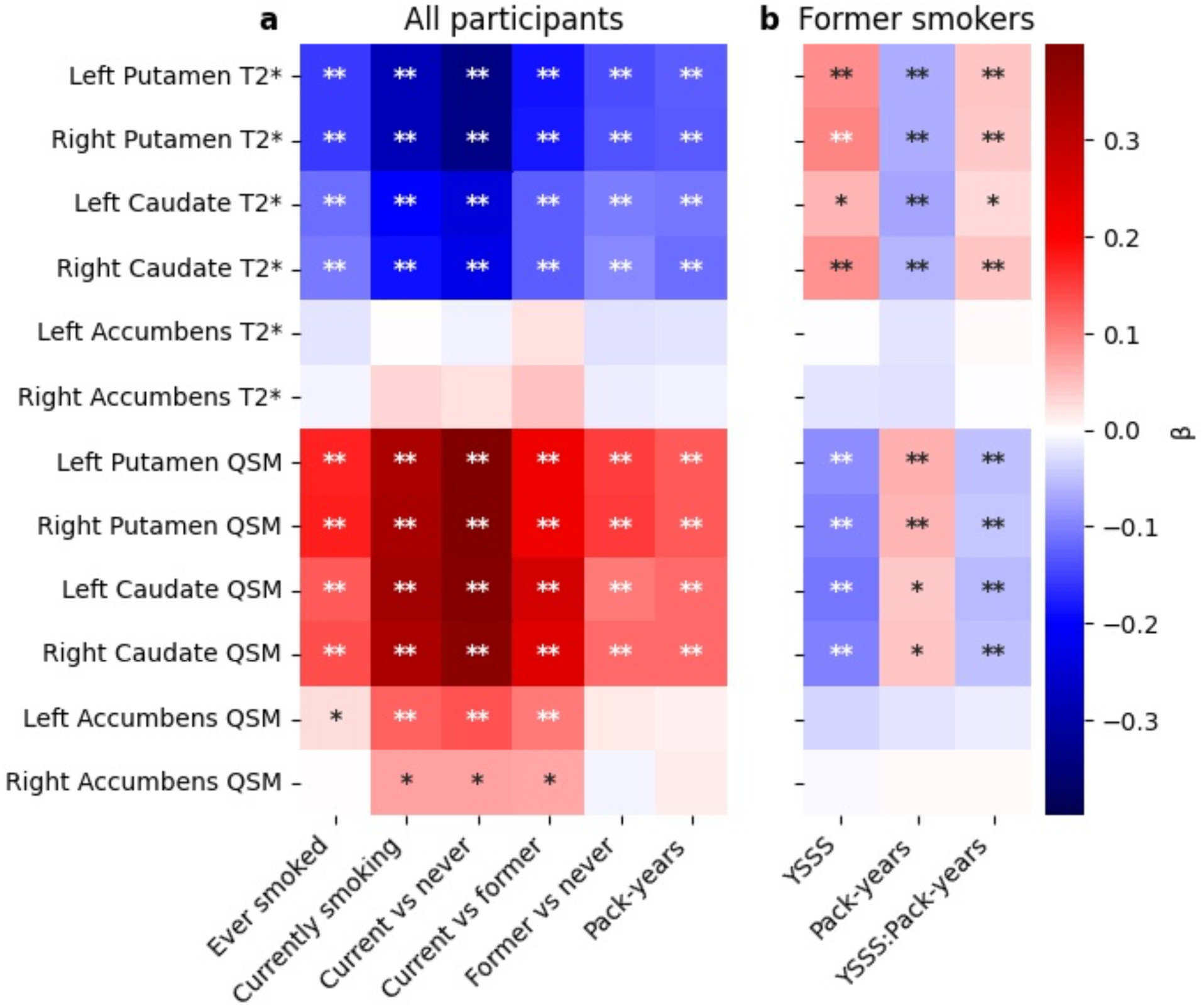
Standardised effect sizes of linear regression models linking striatal iron and smoking in **(a)** the full sample (n≈42k) and **(b)** former smokers (n≈14k). *Lower* T2* and *higher* QSM reflect higher iron content. Prior to performing the regression, IDPs and smoking variables were de-confounded for age, age^2^, sex, age ✕ sex, age^2^ ✕ sex, Townsend deprivation index, income, education, alcohol consumption, blood pressure, BMI, and 25 potential imaging confounders (see Methods). *: FDR-corrected p < 0.05, **: FDR-corrected p < 0.001. QSM: quantitative susceptibility mapping, YSSS: years since stopping smoking.

In the rest of the study, we focused on ‘ever smoked’ since it was the variable for which we found the largest GWAS study with available summary statistics.

### SNP-level genetic correlation

We observed a positive genetic correlation between smoking and QSM in the bilateral putamen and the left accumbens (r ∈ [0.07, 0.14]), and a negative correlation between smoking and T2* in the bilateral putamen (r ∈ [-0.08, -0.07], see the circles in Fig.2). These results were consistent with the corresponding phenotypic correlations (Fig. 2 diamonds, equivalent to Fig. 1a first column but expressed as correlation coefficients rather than effect sizes), with the exception of the caudate where the genetic correlations were no longer significant after correcting for multiple testing. Standard errors were much larger for genetic correlation estimates than for phenotypic correlation estimates (see confidence intervals in Fig. 2 and Supp. Table 5 for details).

**Figure 2.**
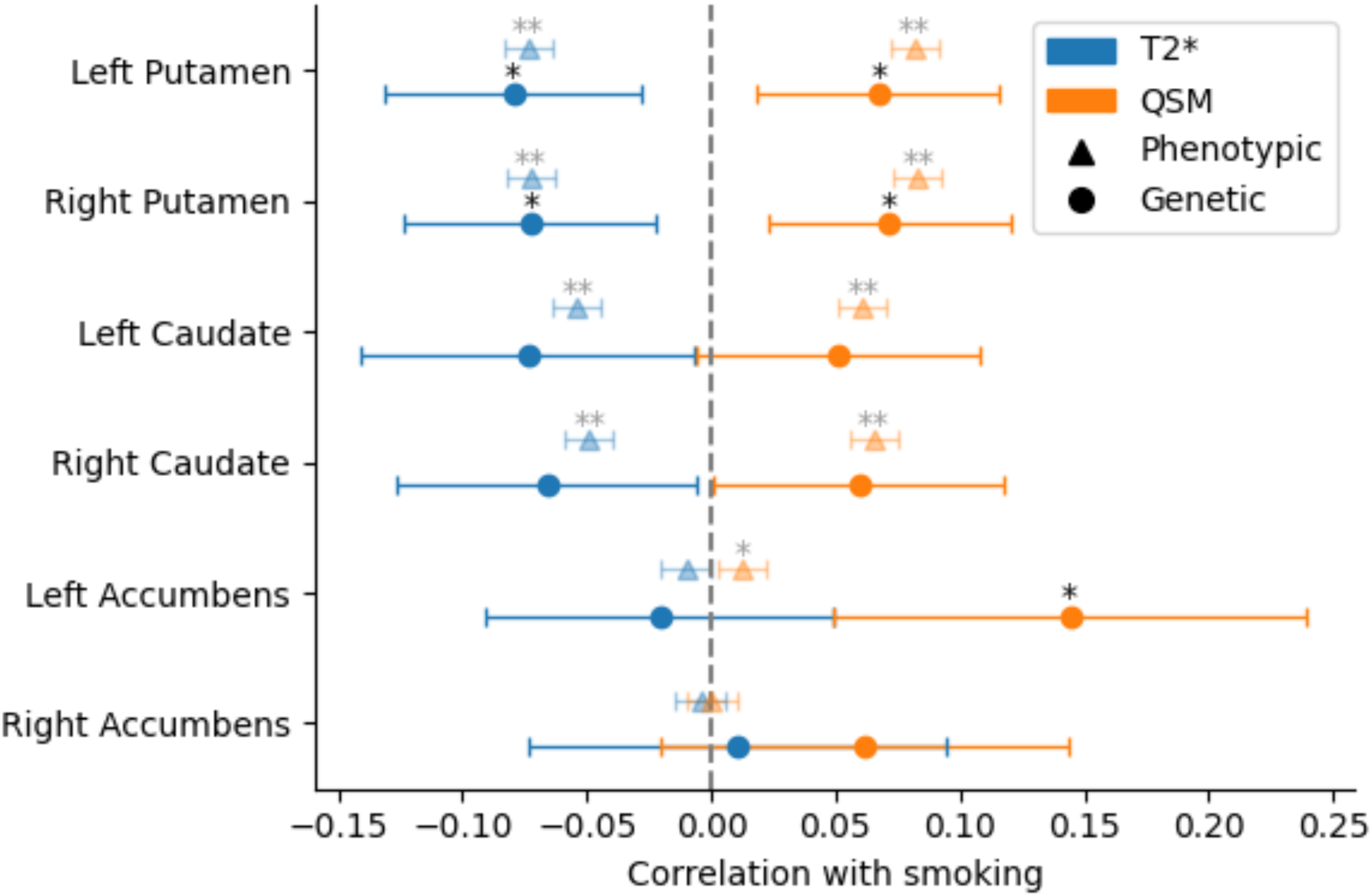
Correlation coefficients between smoking (‘ever smoked’) and striatal iron. Circles represent genetic correlation estimated with LDSR from separate GWAS summary statistics for smoking (n≈500k) and striatal iron IDPs (n≈33k). *Lower* T2* and *higher* QSM reflect higher iron content. Triangles represent phenotypic correlation between the same variables in the UKB (n≈42k) and are shown for comparison (equivalent to Fig. 1 first column but expressed as correlation coefficients rather than betas). Error bars represent 95% confidence intervals. *: FDR-corrected p < 0.05, **: FDR-corrected p < 0.001. QSM: quantitative susceptibility mapping.

### Gene-level analysis

The PascalX cross-GWAS analysis revealed positive correlations between smoking and striatal iron for the dopamine-related genes *DRD2* (in the putamen and caudate) and *PPP1R1B* (all regions, see Fig. 3a). In the exhaustive gene set analysis, *NCAM1* was the only gene found for more than one brain region and contrast (bilateral caudate). Other genes included *DLX5* in the putamen, *CIPC* and *GGACT* in the caudate, and *NAT16*, *NOL4L*, *PLEKHM1* and a gene cluster on chromosome region (chr) 8p23.1 in the accumbens (see Fig. 3b and Supp. Table 6 for detailed results). Those genes are involved in synaptic function and GABAergic and glutamatergic metabolism (52–54).

**Figure 3.**
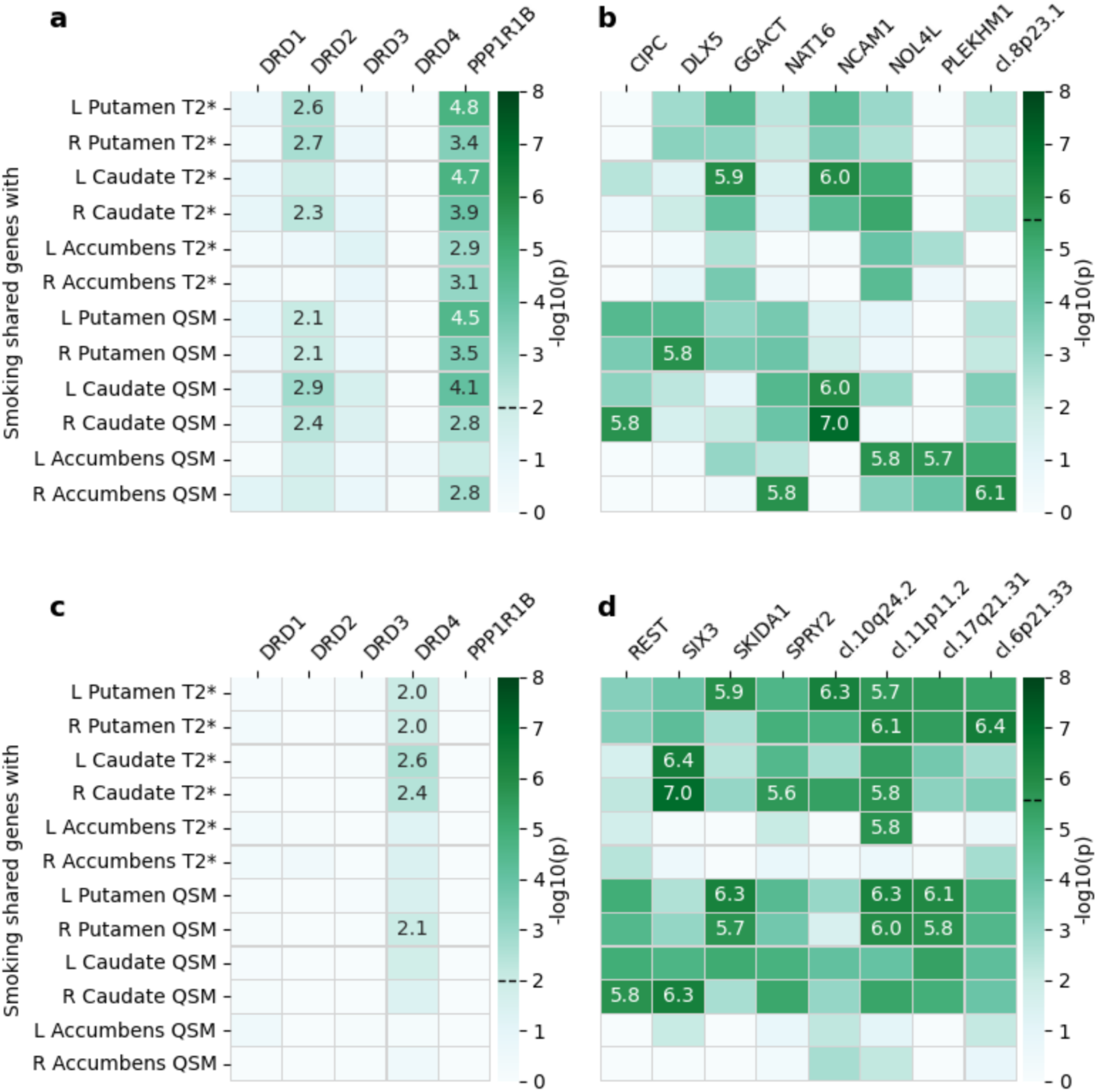
Gene-level correlation between smoking and striatal iron performed using the PascalX cross-GWAS coherence test. We tested **(a, b)** positive and **(c, d)** negative correlations for **(a, c)** five dopamine-related candidate genes and **(b, d)** an exhaustive set of 18 344 genes. –Log10(p) values are annotated for Bonferroni-significant pairs, i.e. with p-values below **(a, c)** 0.01 (0.05/5 candidate genes) and **(b, d)** 2.73×10^-6^ (0.05/18 344 tested genes). Significance thresholds are indicated by dashed lines on the colorbars. L: left, R: right, QSM: quantitative susceptibility mapping. ‘cl.’ indicates gene clusters with their cytogenetic location; cl. 6p21.33: *C6orf25*, *CLIC1*, *DDAH2*, *LY6G6C*, *LY6G6D*, *LY6G6E*, *LY6G6F*, *MSH5*; cl. 8p23.1: *C8orf74*, *PINX1*, *RP1L1*, *SOX7*; cl. 10q24.2: *AS3MT*, *C10orf32*, *CNNM2*; cl. 11p11.2: *C11orf94*, *GYLTL1B*, *PEX16*; cl. 17q21.31: *ARL17B*, *LRRC37A*, *PLEKHM1*.

A negative correlation between smoking and striatal iron content was found for *DRD4* in the putamen and caudate, a gene cluster on chr 11p11.2 (putamen, caudate, and left accumbens), *SKIDA1*, clusters on chr 10q24.2, chr 6p21.33 and chr 17q21.31 in the putamen, and *REST*, *SIX3*, and *SPRY2* in the caudate (Fig. 3c and d and Supp. Table 6). Among other functions, those genes play a role in the immune system, signal transduction, and stress response (54–56).

### Causal relationship

Our MR analysis did not yield significant results. However, there was a trend (p<0.05 that were no longer significant after FDR correction in the bilateral accumbens) suggesting a directional influence from smoking to iron levels rather than from iron levels to smoking (Fig. 4e and 5e, and Supp. Table 7).

**Figure 4.**
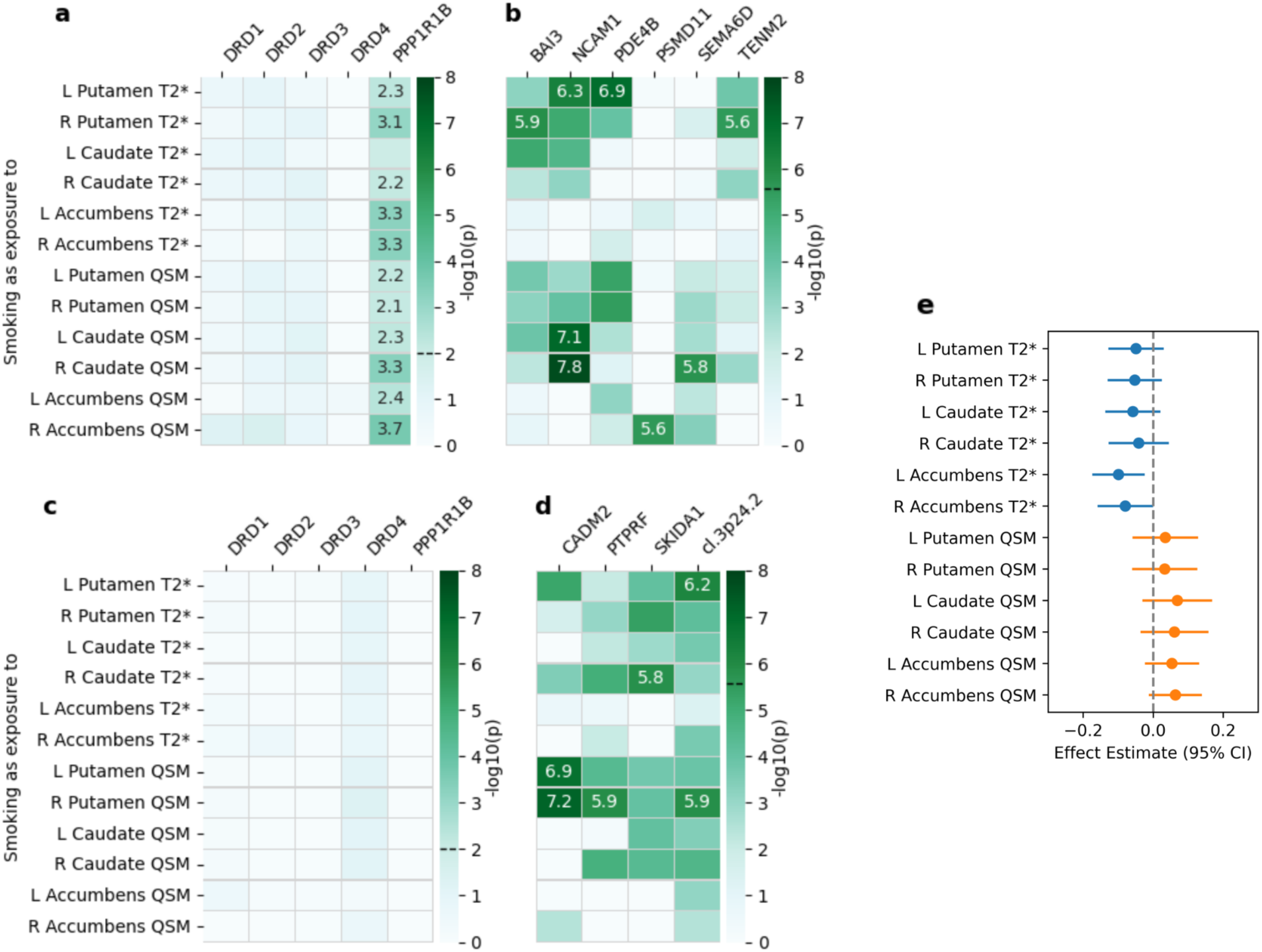
Causality pathway from smoking to striatal iron. For **a-d** we used the PascalX cross-GWAS ratio test for **(a, b)** positive and **(c, d)** negative causal associations in **(a, c)** five dopamine-related candidate genes and **(b, d)** an exhaustive set of 18 344 genes. –Log10(p) values are annotated for Bonferroni-significant pairs, i.e. with p-values below **(a, c)** 0.01 (0.05/5 candidate genes) and **(b, d)** 2.73×10^-6^ (0.05/18 344 tested genes). Significance thresholds are indicated by dashed lines on the colorbars. **(e)** Mendelian randomisation IVW estimates between smoking and striatal iron are indicated with their 95% CI. None of the IVW estimates was significant after FDR correction. CI: confidence interval, IVW: inverse-variance weighted, L: left, R: right, QSM: quantitative susceptibility mapping. ‘cl.’ indicates gene clusters with their cytogenetic location; cl. 3p24.2: *RARB*, *TOP2B*.

**Figure 5.**
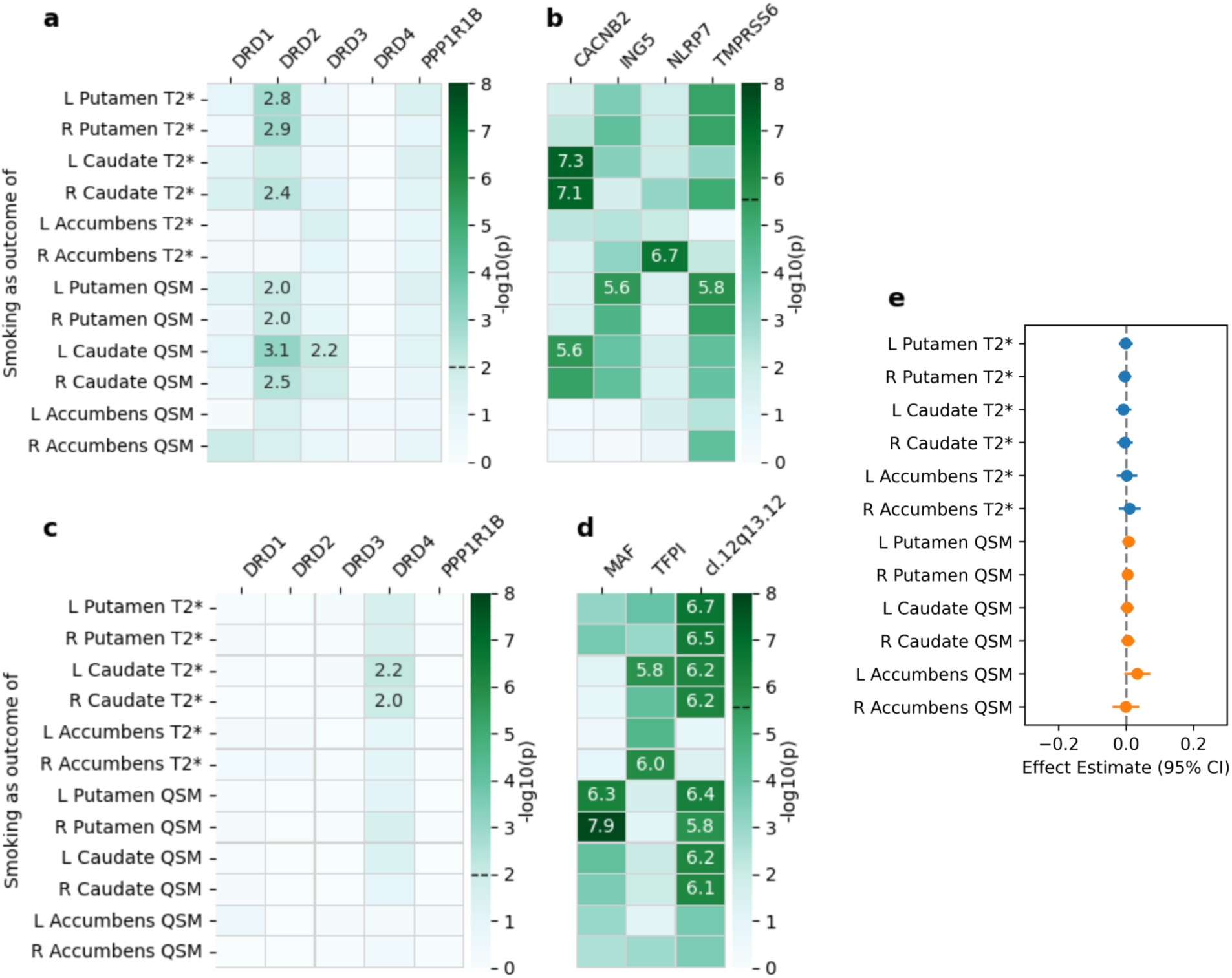
Causality pathway from striatal iron to smoking. For **a-d** we used the PascalX cross-GWAS ratio test for **(a, b)** positive and **(c, d)** negative causal associations in **(a, c)** five dopamine-related candidate genes and **(b, d)** an exhaustive set of 18 344 genes. –Log10(p) values are annotated for Bonferroni-significant pairs, i.e. with p-values below **(a, c)** 0.01 (0.05/5 candidate genes) and **(b, d)** 2.73×10^-6^ (0.05/18 344 tested genes). Significance thresholds are indicated by dashed lines on the colorbars. **(e)** Mendelian randomisation IVW estimates between striatal iron and smoking are indicated with and their 95% CI. None of the IVW estimates was significant. CI: confidence interval, IVW: inverse-variance weighted, L: left, R: right, QSM: quantitative susceptibility mapping. ‘cl.’ indicates gene clusters with their cytogenetic location; cl. 12q13.12: *METTL7A*, *TFCP2*.

The PascalX ratio test indicated positive bidirectional causality through genes expressed in brain tissues (detailed results in Supp. Tables 8 and 9). Specifically, for the pathway from smoking to striatal iron, the implicated genes were *PPP1R1B*, *BAI3*, *NCAM1*, *PDE4B*, *PSMD11*, *SEMA6D*, and *TENM2* (Fig. 4a and b), which are associated with dopaminergic and glutamatergic activity, synaptic plasticity, and neuroinflammation (54,57–59). Conversely, for the pathway from striatal iron levels to smoking, the involved genes were *DRD2*, *DRD3*, *CACNB2*, *ING5*, *NLRP7*, and *TMPRSS6* (Fig. 5a and b), linked to dopamine signalling, iron homeostasis, and inflammation (54,60,61).

Additionally, the PascalX ratio test identified negative causality (i.e. between smoking and *lower* iron). For the pathway from smoking to iron content in the putamen and caudate, the genes *CADM2*, *PTPRF*, *SKIDA1*, and a gene cluster on chr 3p24.2 (Fig. 4c and d) were involved, primarily associated with immune system functions and synapse organisation (54,62,63). For the reverse pathway, from striatal iron to smoking, the genes *DRD4*, *MAF*, *TFPI*, and a gene cluster on chr 12q13.12 were implicated (Fig. 5c and d), with functions related to dopamine signalling, inflammatory response, and the immune system (54,64,65).

## Discussion

In this study, we identified a positive association between tobacco smoking and increased iron content in the striatum, particularly in the dorsal striatum (putamen and caudate). Notably, iron accumulation in the dorsal striatum was positively correlated with pack-years of smoking and negatively correlated with the number of years since smoking cessation, with stronger effects observed in individuals with greater lifetime tobacco exposure. These findings could not be attributed to linear effects of alcohol consumption, blood pressure, BMI, demographic, socio- economic, or imaging variables, as we corrected for these confounders prior to the analysis.

Additionally, we observed a global positive genetic correlation between smoking and iron levels in the putamen and left accumbens. Gene-specific analyses revealed a complex pattern of coherent and anti-coherent associations between smoking and iron, reflecting diverse genetic pathways. Evidence of bi-directional causal relationships was found, involving genes related to synaptic function (*BAI3*, *CACNB2*, *CADM2*, *NAT16*, *NCAM1*, *PLEKHM1*, *PTPRF*, *SEMA6D*, *TENM2*), dopaminergic (*DRD2*, *DRD3*, *DRD4*, *NCAM1*, *PPP1R1B*), GABAergic (*DLX5*), and glutamatergic (*GGACT*, *PPP1R1B*) transmission, as well as immune function, inflammation, and stress response (*NLRP7*, *MAF*, *METTL7A*, *REST*, *SIX3*, *SKIDA1*, *SPRY2*, *TFCP2*, *TOP2B*, and a gene cluster on chr 6p21.33).

The association of iron levels with smoking in the dorsal striatum appears to be cumulative, as it is proportional to pack-years, and potentially reversible, given that individuals with a longer history of smoking cessation showed lower iron levels. Although this is speculative given the correlational nature of our results and would require confirmation in a longitudinal design, if true, it suggests that smoking cessation could have a positive impact on brain health, similar to the gradual recovery of lung function observed after quitting smoking (66). In contrast, iron levels in the ventral striatum (accumbens) were elevated only in current smokers and showed no correlation with pack-years. This might be due to the differential dopamine signalling responses to nicotine between the ventral and dorsal striatum, limiting long-term effects primarily to the dorsal striatum (67). Research indicates that the ventral striatum initially drives voluntary drug use, while the dorsal striatum increasingly takes over as habitual patterns emerge, facilitating the progression to compulsive behaviour during habit formation (68). The leftward asymmetry that we observed in the accumbens could be interpreted in light of disrupted accumbens volume asymmetry reported in a recent multi-cohort study, where nicotine-dependent individuals had larger left accumbens volumes than non-dependent controls (69). The fact that we observed a link between smoking and left accumbens iron when using QSM, but not T2*, as an iron marker could indicate concurrent effects on myelin content.

Iron accumulation in the brain may be linked to ferroptosis, an iron-dependent type of cell death that is accompanied by iron deposition and glutamate toxicity (20,70). Ferroptosis has also been linked to inflammatory pathways (71). Our identification of genes involved in oxidative stress, inflammation, and immune response aligns with the hypothesis of heightened ferroptosis and inflammation related to smoking. This finding is particularly important given that smoking is a known risk factor for Alzheimer’s disease and other neurodegenerative diseases, where oxidative stress and neuroinflammation have been specifically identified as key mechanisms (14,22,23).

Mendelian randomisation (MR), a well established tool to investigate causality, did not produce significant results, although there was a trend indicating that smoking influences iron levels rather than vice versa. This trend aligns with previous research linking substance dependence to brain MRI measures (6,10,27). We hypothesise that the lack of significant results from MR may stem from the complexity of smoking and brain iron as polygenic traits, which are influenced by numerous factors. This complexity can make it challenging to identify causal relationships using a limited number of SNPs as genetic instruments. Indeed, significant findings emerged when aggregating SNP-wise signals at the gene-level using our recently introduced PascalX methodology. Consistent with the observed trend in MR, we identified more genes with smoking as exposure and iron as outcome than vice versa. Moreover, the ability of PascalX to distinguish between coherent and anti-coherent gene signals, may explain its power to detect causal relationships despite the lack of significant MR results, as such signals would largely cancel each other in the latter. As we discuss in the following, the vast majority of genes detected with PascalX appear functionally plausible. This indicates that PascalX can be a powerful extension of standard MR, in particular when results are inconclusive or only marginally significant.

We hypothesised that the relationship between smoking and striatal iron would involve the dopaminergic system. Two dopamine receptor genes, *DRD2* and *DRD4*, were identified with opposing effects in the dorsal striatum. *DRD2* (along with *DRD3* in the causal analysis) was genetically associated with smoking and higher iron levels in the dorsal striatum, whereas *DRD4* was linked to lower iron. Both genes were implicated in the causal pathway from striatal iron to smoking, suggesting that an imbalance in striatal dopamine receptors may increase the likelihood of initiating smoking. *PPP1R1B*, a gene that regulates dopaminergic and glutamatergic signalling and plasticity in the striatum, showed coherent signals for smoking and iron content, with a causal direction from smoking to iron accumulation. A similar pattern was observed for *NCAM1* in the dorsal striatum. *NCAM1* has previously been identified as a binding partner of *DRD2* and a modulator of dopaminergic activity, forming a complex with *DRD2* upon dopamine stimulation, particularly through its *NCAM180* isoform (72). The *NCAM1* gene is part of a cluster (*NCAM1-TTC12-ANKK1-DRD2*) implicated in various dopamine-related disorders, including attention-deficit/hyperactivity disorder and substance dependence (73–75). Our findings expand this understanding by suggesting a causal relationship in which smoking behaviour leads to iron accumulation in the striatum via stimulation of the dopaminergic system.

Beyond genes involved in dopamine transmission, we identified several genes linked more broadly to synaptic function, predominantly in the causal pathway from smoking to dorsal striatal iron. This suggests that synaptic plasticity in smokers may contribute to increased iron content in the dorsal striatum. Alternatively, specific synaptic organisation during development could predispose individuals to smoking, with iron accumulation occurring as a downstream effect.

Several genes previously associated with substance abuse beyond cigarette smoking (*BAI3*, *CADM2*, *ING5*, *PSMD11*) (76–78) and, more generally, with impulsive behaviour and risk-taking (*BAI3*, *CADM2*, *PTPRF*) (79) point to potential mechanisms, such as synaptogenesis and signalling (*BAI3*, *CADM2*) (78,80), increased acetylation of histones H3 and H4 within the reward circuitry (*ING5*) (81), and protein-protein interactions at synapses (*PTPRF*) (62).

Our study has several limitations. The phenotypic analysis was conducted primarily in samples of White European ancestry, and the genetic analysis was limited exclusively to European ancestry, which restricts the generalisability of our findings. Although a recent multi-ancestry smoking GWAS has been published (82), there is currently no large-scale GWAS of brain iron markers in diverse populations, which is needed to replicate our findings across different ancestries. Ideally, the genetic analysis would be validated in an independent cohort, but this was not feasible since the UKB is the only study of sufficient size that includes brain swMRI data. Additionally, while we controlled for potential confounders in the phenotypic analysis, the same adjustments were not possible in analyses relying on GWAS summary statistics from previous studies. Therefore, confounding effects—particularly from alcohol consumption, which is linked to elevated striatal iron (27,83,84)—cannot be fully ruled out in the genetic findings.

In summary, our study reveals a complex relationship between smoking and striatal iron levels, with overall positive genetic and phenotypic correlations. Gene-level analyses suggest a trend where smoking primarily influences iron levels, although both positive and negative associations were observed, indicating intricate bidirectional mechanisms and possible feedback loops. Our findings suggest that smoking may lead to increased striatal iron accumulation through multiple pathways, which may impact brain health and neurodegenerative risk. Additionally, inflammation and dopamine imbalance in the striatum may further reinforce smoking behaviour. Further research, particularly involving more diverse populations and longitudinal data, is essential to fully elucidate these dynamics.

## Data and code availability

UKB data are available upon successful application (https://www.ukbiobank.ac.uk/enable-your-research/apply-for-access). GWAS summary statistics used in this study are publicly available at https://www.fmrib.ox.ac.uk/ukbiobank/gwas_resources/ and https://open.win.ox.ac.uk/ukbiobank/big40/ for brain IDPs and https://conservancy.umn.edu/bitstream/handle/11299/201564/SmokingInitiation.txt.gz for smoking initiation. The code used to generate the presented results will be made available on GitHub upon publication (https://github.com/ot710/smoking_striatum_iron).

## Acknowledgements

This research has been conducted using the UK Biobank Resource under Application Number 90947. The authors thank Sofía Ortín Vela, Dennis Bontempi, Michael Beyeler, and David Presby for their valuable feedback and insightful suggestions on the manuscript.

