## Supplementary Figures for "Bidirectional genetic and phenotypic links between smoking and striatal iron content involving dopaminergic and inflammatory pathways"

**Supplementary Figure 1.** T2* and QSM distributions by sex. Male and female means were compared using two-sample t-tests. Respective means are indicated along with effect sizes (Cohen's d) when the p-value was <0.05. Male and female variances were compared using F-tests. F mean: female mean, M mean: male mean, QSM: quantitative susceptibility mapping.

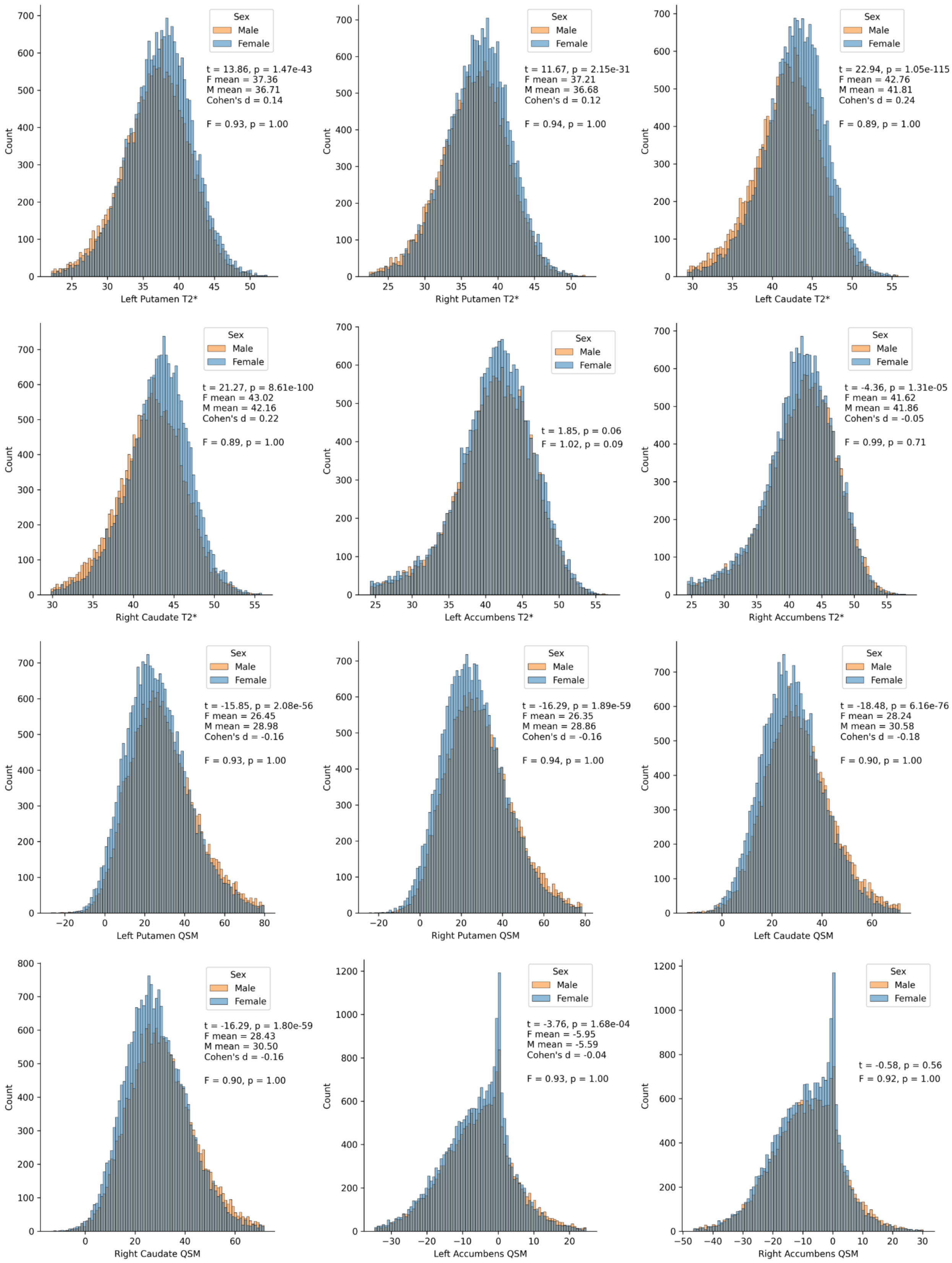

Sex
Male
Female
t = 13.86, p = 1.47e-43
F mean = 37.36
M mean = 36.71
Cohen's d = 0.14
F = 0.93, p = 1.00
Count
Left Putamen T2*
t = 11.67, p = 2.15e-31
F mean = 37.21
M mean = 36.68
Cohen's d = 0.12
F = 0.94, p = 1.00
Right Putamen T2*
t = 22.94, p = 1.05e-115
F mean = 42.76
M mean = 41.81
Cohen's d = 0.24
F = 0.89, p = 1.00
Left Caudate T2*
t = 21.27, p = 8.61e-100
F mean = 43.02
M mean = 42.16
Cohen's d = 0.22
F = 0.89, p = 1.00
Right Caudate T2*
t = 1.85, p = 0.06
F = 1.02, p = 0.09
Left Accumbens T2*
t = -4.36, p = 1.31e-05
F mean = 41.62
M mean = 41.86
Cohen's d = -0.05
F = 0.99, p = 0.71
Right Accumbens T2*
t = -15.85, p = 2.08e-56
F mean = 26.45
M mean = 28.98
Cohen's d = -0.16
F = 0.93, p = 1.00
Left Putamen QSM
t = -16.29, p = 1.89e-59
F mean = 26.35
M mean = 28.86
Cohen's d = -0.16
F = 0.94, p = 1.00
Right Putamen QSM
t = -18.48, p = 6.16e-76
F mean = 28.24
M mean = 30.58
Cohen's d = -0.18
F = 0.90, p = 1.00
Left Caudate QSM
t = -16.29, p = 1.80e-59
F mean = 28.43
M mean = 30.50
Cohen's d = -0.16
F = 0.90, p = 1.00
Right Caudate QSM
t = -3.76, p = 1.68e-04
F mean = -5.95
M mean = -5.59
Cohen's d = -0.04
F = 0.93, p = 1.00
Left Accumbens QSM
t = -0.58, p = 0.56
F = 0.92, p = 1.00
Right Accumbens QSM

**Supplementary Figure 2.** T2* and QSM values by age. Pearson's correlation coefficients and p-values are indicated. QSM: quantitative susceptibility mapping.

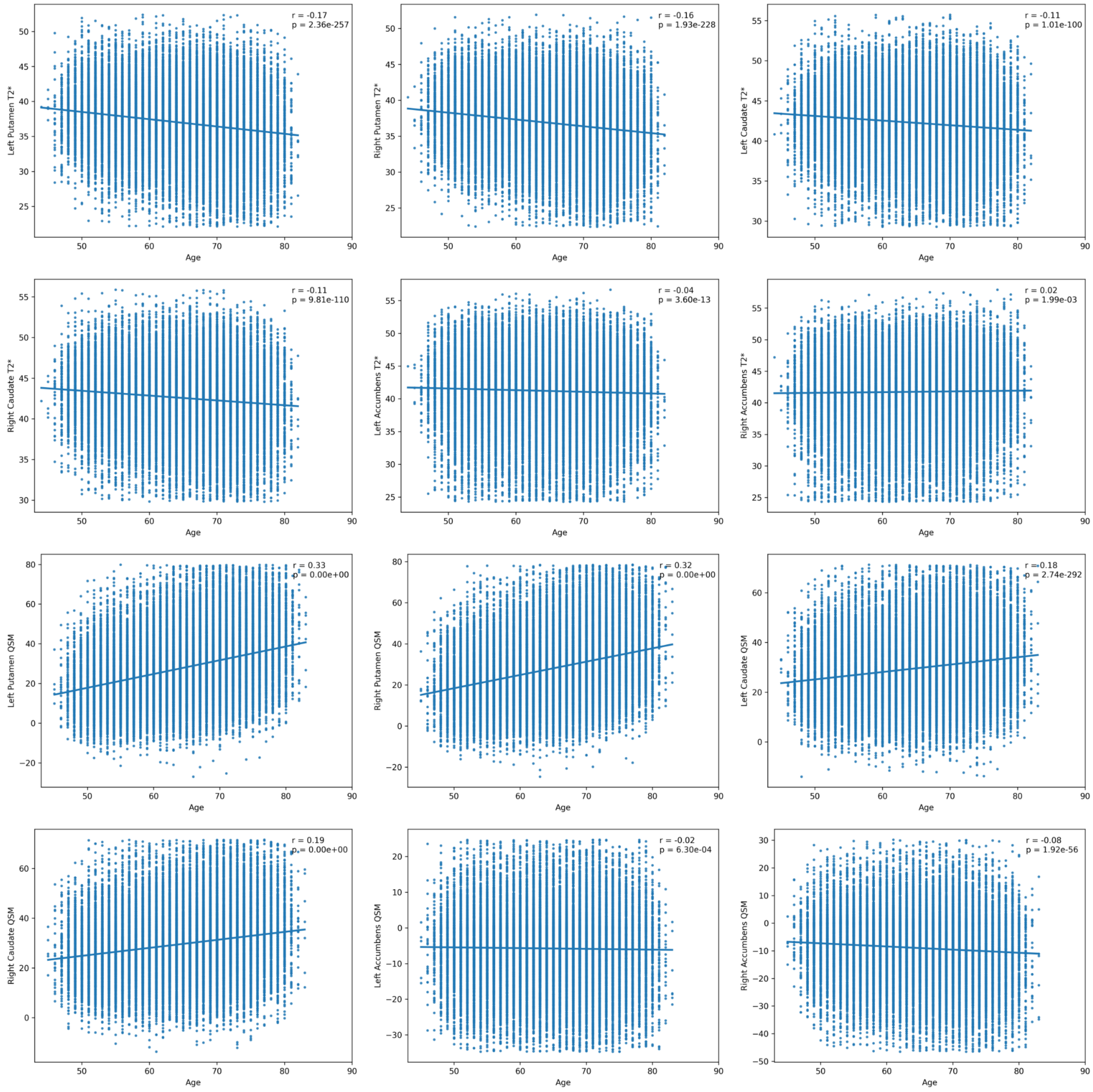

**Supplementary Figure 3.** T2* and QSM distributions by smoking status and sex. No statistical tests were used to compare the groups. QSM: quantitative susceptibility mapping.

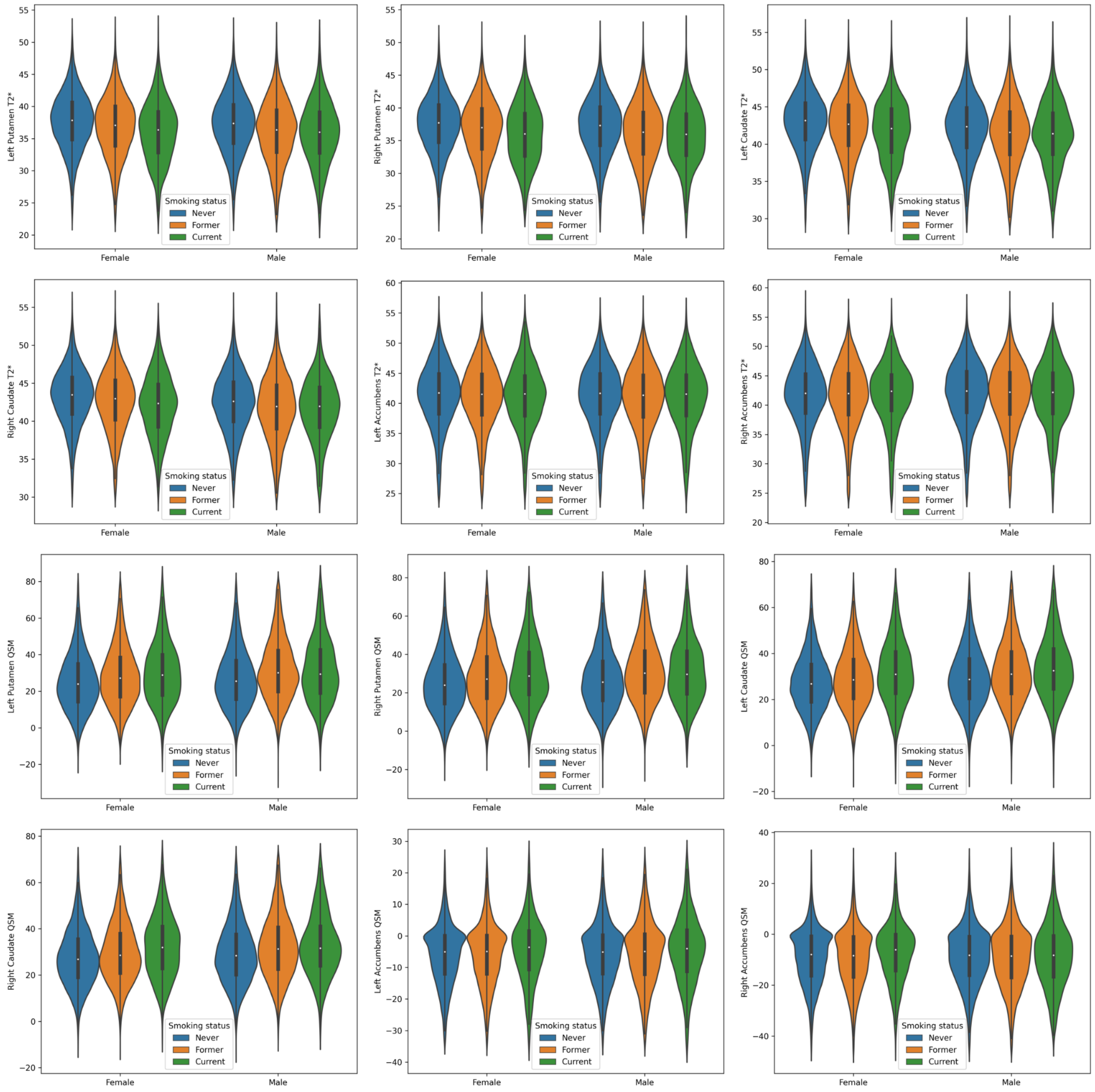

**Supplementary Figure 4.** Beta coefficients of smoking-by-sex and smoking-by-age interaction terms in linear regression models linking striatal iron and smoking. Linear models also included main effects of smoking and sex, respectively age, but only the interaction effects are shown here. None of them was statistically significant. QSM: quantitative susceptibility mapping.

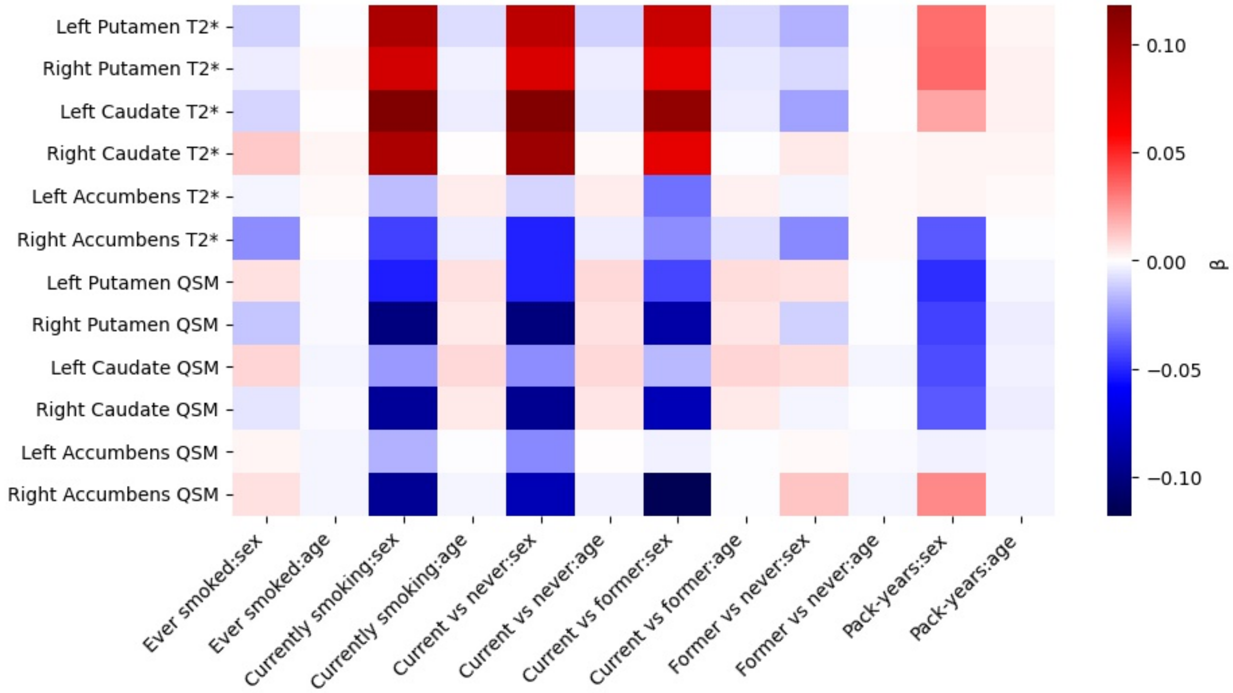

**Supplementary Figure 5.** Right putamen QSM in former smokers by YSSS and pack-years quartiles. We show the right putamen QSM as an example, since a similar pattern was observed in all dorsal striatum regions (see Fig. 1b). The dashed line indicates the mean value of never smokers, while the dotted line, that of current smokers in the fourth pack-years quartile. Coloured lines represent regression lines of respective pack-years quartiles in former smokers. The interaction between YSSS and pack-years is made visible by the difference in slopes between quartiles–while the first quartile's QSM mean is very close to that of never smokers regardless of YSSS, the QSM mean of the fourth quartile is higher at 0 YSSS and similar to never smokers at about 40 YSSS. CS: current smokers, NS: never smokers, QSM: quantitative susceptibility mapping, YSSS: years since stopping smoking.

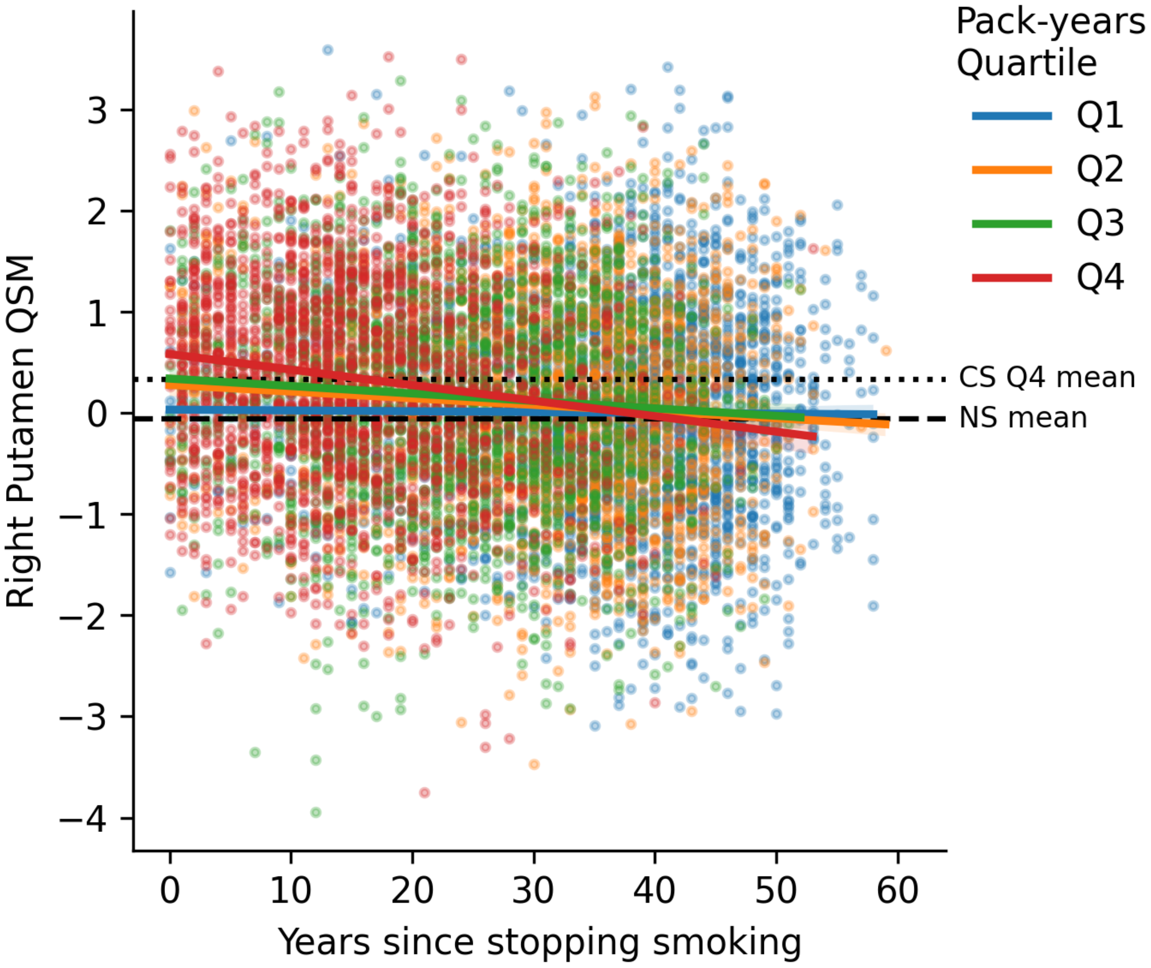
